# Targeted Interventions to Improve Discharge Times in an Australian Cardiology Department: a Before-and-After Study

**DOI:** 10.1101/2025.05.21.25328063

**Authors:** Eddy Xiong, Dilki Da Silva, Mel Elsing, Donna Perry, Mitch Eldridge, Angela Carberry, Virginia Decourcy, Zachary King, Jared Pleash, Anneline Helms, Kim Greaves

## Abstract

**Background:** Late discharges in cardiology inpatient units contribute to patient flow inefficiencies, emergency department overcrowding, and reduced hospital capacity. We define late discharges as those that occur after 12:00pm.

**Methods:** We conducted a quality improvement initiative using the Plan-Do-Check-Act framework over the course of one year. We evaluated a baseline mean discharge time in patients over the course of one month and identified causes of delays. Then, through iterative qualitative analysis we generated and implemented new departmental policies aimed at improving discharge times. We then re-assessed discharge outcomes at one, and six months post intervention.

**Results:** Following intervention, we saw a reduction in median discharge time of 1h 27m (95% CI 0:11-2:14, p<0.01) after one month compared to baseline. This was sustained at after six months with a 1h 20m reduction (95% CI 0:23-2:10, p<0.01), with no significant difference between the one and six month periods. Participants were also more likely to have a timely discharge before 12:00 at one (RR=3.03, 95% CI 1.84 – 4.99. p<0.01) and six (RR=2.39, 95% CI 1.43 – 4.01. p<0.01) months, compared to baseline.

**Conclusion:** Targeted interventions significantly improved discharge efficiency in a cardiology inpatient unit, enhancing patient flow and operational performance. This framework can inform similar initiatives in other settings.

## Introduction

An aging global population has accelerated healthcare needs manifesting in an increase in inpatient hospital admissions, with significant concerns that if left unchecked, demand may outstrip existing hospital capacities [1]. This is particularly prevalent in Australia, where decreased bulking billing rates and higher gap payments for primary care have resulted in patients forgoing regular GP preventative care, often meaning hospital visits are more likely to result in admission [2].

Definitions for ‘early discharge’ vary depending on local policy, however for the purposes of this study we define it as discharges occurring before 12:00pm [3]. The importance of timely discharge should not be understated. Late discharges, which often occur in the afternoon, have a significant impact on patient flow throughout the hospital. Emergency department presentations in Australia typically surge at noon, with a gradual decline in the afternoon [4]. This makes early morning discharges of paramount importance to prevent emergency department bottlenecks and avoid adverse outcomes. A major hindrance to efficient inpatient admissions is an inefficient discharge system, as this can result in a back log of patients unable to be admitted and subsequent ‘bed blocking’. The outcome of this is significant overcrowding of the Emergency department leading to consequences such as ambulance ramping, subsequently impacting on key outcomes such as length of stay, triage time, and admission rates [5, 6]. Subsequently, this has been demonstrated to result in significant preventable illness and death, often secondary to issues such as delayed care and increased exposure to hospital acquired pathogens [7, 8]. Additionally, some studies have suggested a direct correlation between delayed ambulance offloading times and increased patient mortality [9].

The reasons for delays in discharge are not always immediately apparent [10]. It is highly multifactorial and often arises from a mixture of patient factors and organisational factors related to healthcare and hospital management, which makes it a difficult issue to address. Whilst complex patient needs can complicate and hinder the discharge process, this is often compounded by inefficiencies in the discharge processes facilitated by hospital staff [11]. Common causes of delays related to healthcare included waiting for health evaluations, treatments, exam results, and discharge preparations [12-14]. Various methods of ameliorating these causes have been implemented in other studies such as earlier review of patients suitable for discharge, however often barriers to discharge are highly contextual and specific to individual health services [15]. Furthermore, existing evidence suggests that merely asking clinicians to prioritise seeing early discharges led to no change in mean discharge time or length of stay. As such, additional measures may be necessary to result in early discharge[16]. Previous studies addressing this issue have approached the issue using a Plan-Do-Check-Act model to produce tailored guidelines with successful outcomes [17, 18].

Whilst definitions for ‘patient discharge’ can vary, for the purposes of this study we define it as a patient leaving the inpatient ward allowing their bed to become available for another patient. This can involve being transferred to a transit lounge, which is a dedicated area in the hospital capable of temporarily holding patients either suitable for discharge or presenting from the emergency department requiring an inpatient bed.

This study sought to understand the barriers to and enablers of earlier discharge in cardiology inpatients of an Australian tertiary hospital. Applying the Plan-Do-Check-Act framework, we then used this information to co-design solutions to overcome these issues, implemented them and measured their effectiveness in improving discharge times, both in the short and longer terms. Whilst there is existing research regarding this topic, to our knowledge this is the first study to address this issue in the context of a Cardiology department.

## Methods

Our study was a quality improvement activity in the setting of a cardiology department at the Sunshine Coast University Hospital in Queensland, Australia, in which we drafted and implemented a set of operational guidelines with the goal of improving discharge times. The 24-bed cardiology department facilitates all tertiary services except for cardiothoracic surgery and has a full suite of interventional suites including electrophysiology and primary percutaneous coronary angiography. It services a Modified Monash category 1 region and receives patients from five smaller peripheral hospitals [19]. For the year 2024, the department had a total of 1891 inpatient admissions. Monthly acute patient admissions were consistent, averaging 160 patients. The study was conducted over the duration of six months, from February to November 2024 however the intervention remains in place at time of publication.

The project team included a Cardiology Consultant, three Cardiology Advanced Trainees, a Resident Medical Officer, one medical student, three nursing unit managers, and two research officers. The B410 discharge study was conducted in three broad phases, applying the Plan-Do-Check-Act model (Figure 1).

**Figure 1.**
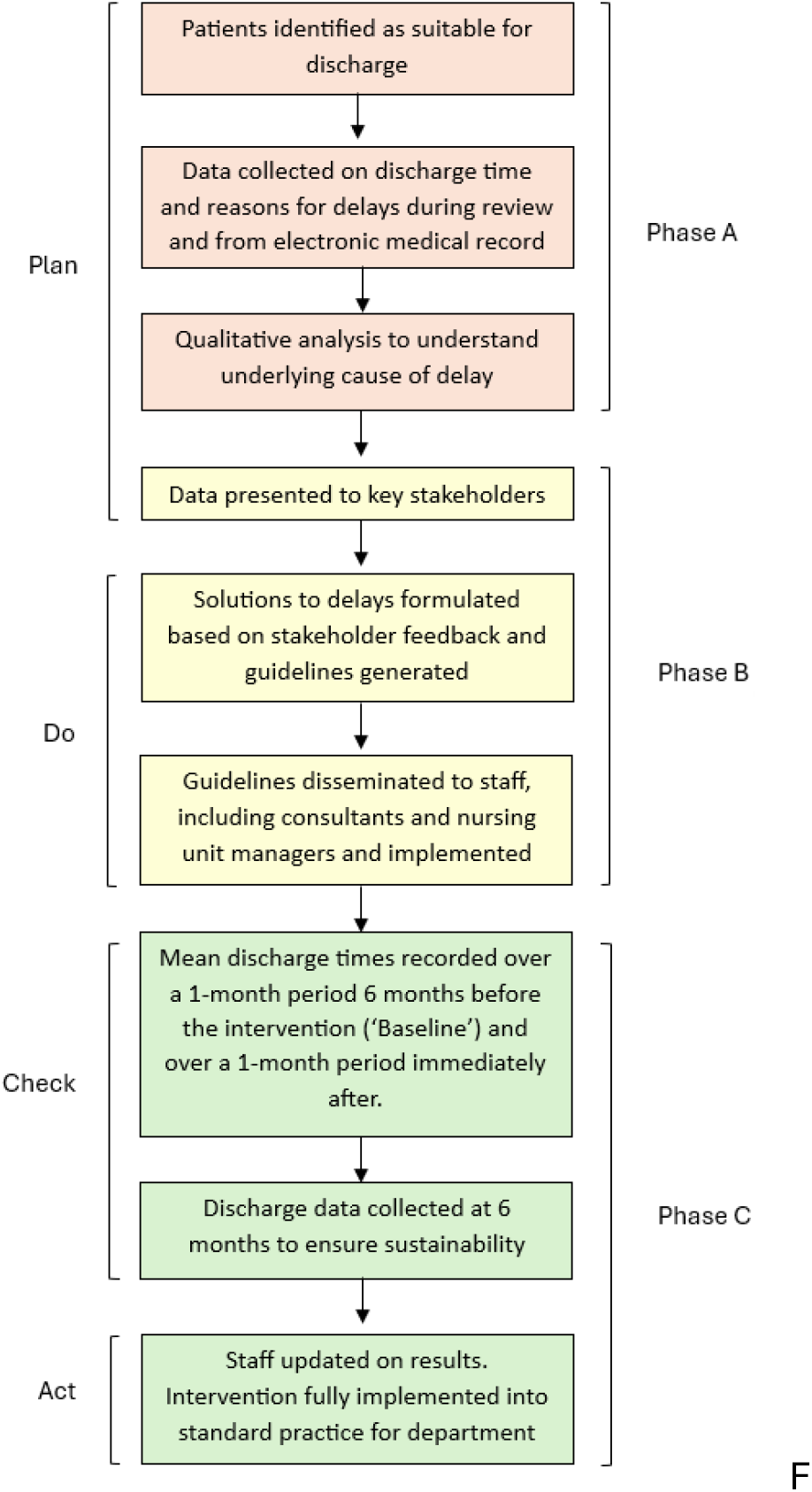
Flow chart representing phases of B410 discharge study.

### Phase A: Baseline assessment of discharge times and reasons for delays

Phase A of the project was conducted over a 5-day period excluding the weekend in May 2024. The project team met each afternoon and identified patients likely to be discharged the following day, kept a record of the times patients were subsequently discharged, any delays, and the reason(s) for the delay. This information was documented on a collaborative spreadsheet by various team members and then corroborated with the patient’s electronic medical record.

Quantitative categorical variables for discharge time (before 10:00, 10:00-12:00, after 12:00) and reason for discharge delay were examined. We identified primary and secondary cause of delayed discharge. We define a primary cause of delay as the issue or event which directly resulted in the longest delay in discharge. Secondary causes of delay refer to those which resulted in a significant delay (more than two hours) despite not being the primary cause.

Integrated electronic medical record notes were examined and anecdotal evidence from nursing and medical staff was also collected. These data were then categorised to examine underlying drivers of delays.

### Phase B: Development and Implementation of Guidelines

Insights from Phase A were presented to a group of key stakeholders in May including cardiologists, clinical nurses, and junior doctors to formulate potential solutions as well as gain additional perspectives on reasons for delays. A set of draft guidelines was generated and then presented to cardiology staff for initial feedback and review which then guided the development of revised guidelines. Implementation began later that month, when the guidelines were disseminated amongst staff and presented at discipline-specific meetings. Daily check-ins and weekly stakeholder meetings were undertaken to collect feedback and inform adjustments to the guidelines.

### Phase C: Evaluation and Statistical Analysis

Data on discharge times was collected and a comparative quantitative analysis conducted to evaluate the impact of the guidelines. Data regarding discharge times for each patient in the cardiology ward was collected for three one month periods: baseline - six months before intervention (November 2023), conducted well before the conception of the study to minimise potential bias; one month following intervention (27 May – 21 June 2024, weekends excluded) and six months following intervention (1 – 30 November 2024, weekends excluded).

Median discharge time was compared between the second group and baseline, between the third group and baseline, and between the two intervention groups. Categorical variables for discharge time (before 10:00, 10:00-12:00, after 12:00) were also created and compared between groups. Statistical significance was set at p<0.05. Appropriate tests, including the Mann-Whitney U test, Fisher’s exact test, and Chi-squared test, were used to calculate p values based on the type of data. We used a kernel density estimation within the R software application to generate a density plot comparing the results of the three periods.

#### Ethics

This evaluation was classified as a quality improvement initiative and thus an ethics exemption was granted from Gold Coast Hospital Health Service Human Research Ethics Committee (HREC) - EX/2025/QGC/116865

## Results

### Phase A

During Phase A, over the period of one week we identified 38 inpatients who were suitable for discharge the following day. Of these, 20 were suitable for early discharge. These patients were used to identify potential reasons for delays in discharge. Of these, one patient was discharged prior to 10:00, 3 were discharged between 10:00-12:00, and 16 were discharged after 12:00. For the 16 patients discharged after 12:00, reasons for delays were identified and grouped (table 1).

**Table 1.** Reasons for delayed discharge (After 12:00) for inpatients in a cardiology unit, collected over one week (May 2024).

| Reasons for later discharge | Primary cause (%) | Secondary cause <sup>†</sup> (%) | Total |
| --- | --- | --- | --- |
| Delayed ward round | 4 (25.0) | 1 (6.3) | 5 |
| Awaiting discharge paperwork | 3 (18.8) | 0 | 3 |
| Awaiting review from another team* | 2 (12.5) |  | 2 |
| Awaiting transport home | 2 (12.5) | 0 | 2 |
| Pharmacy review | 2 (12.5) | 2 (12.5) | 4 |
| Other causes |  |  |  |
| Awaiting Chest X-Ray | 1 (6.3) | 0 | 1 |
| Awaiting echocardiogram report | 1 (6.3) | 0 | 1 |
| Awaiting ambulance transfer | 1 (6.3) | 0 | 1 |

\*Other teams included- social work, indigenous health liaison officer, clinical nurse review, other medical speciality e.g. Neurology.
†Primary cause describes the most significant factor resulting in late discharge, and secondary cause refers to any other contributing cause.

Patients sometimes had multiple contributing factors to their delay, as one late review can lead to a cascading effect where subsequent reviews or patient needs are then delayed. The most common primary reasons were a delay in ward round, or the patient being provided discharge paperwork. Another major reason for delay was if patients were suitable for discharge, however, were awaiting a family member or friend to pick them up.

Five broad causes of delayed discharge were categorised and summarised as follows:

#### 1. Delayed Ward round

Various factors contributed to patients being seen late on the morning ward round. The most common cause was that patients with higher clinical urgency took priority being seen. Other causes included delayed morning meetings, often secondary to lengthy handovers. Consultant meetings, held after the morning handover also contributed as the ward round was unable to commence until the rounding cardiologist arrived, compounded by a lack of urgency by the consultant to commence the ward round. Feedback from resident medical officers and nursing staff indicated that restructuring the department’s morning schedule would likely be necessary to address this issue.

#### 2. Required imaging

Patients requiring ‘discharge-dependent’ imaging such as a chest X-ray after a pacemaker insertion the previous day, or an echocardiogram were very rarely discharged before 12:00. Staff observed that the delay was largely organisational and occurred at multiple levels. Such patients needed to be first seen on the morning ward round, then transported to radiology, the results reviewed by the treating team, and a clinical decision made regarding discharge.

#### 3. Requiring review by other disciplines (allied health) before discharge

Review by another discipline before discharge such as pharmacy or cardiac rehabilitation, was often a final step in the process which delayed patient departure times. The reasons for this were because other disciplines had other priorities or were unaware of the department’s goal of timely discharge.

#### 4. Delayed paperwork provision

Despite being seen relatively early on ward rounds, patients did not receive their discharge paperwork in a timely fashion and as such their discharge was delayed. This was because resident medical officers had other competing ward duties. Input from residents suggested this mainly occurred in the context of delayed ward rounds which were often rushed to make up for the lost time. This meant that discharge summaries could only be completed after the round.

#### 5. Awaiting transport

Some patients, despite being fit for discharge before noon, were unable to leave until late in the afternoon due to waiting for transport by family member or friend. Multiple staff members identified that these patients were potentially suitable to being discharged to the hospital’s transit lounge, thereby freeing up a hospital bed.

### Phase B

Following presentation of Phase A findings to key stakeholders, we developed a set of key themes and guidelines for improving patient flow and discharge times:

1. Optimizing the Use of the Transit Lounge
  - Patients identified for discharge should be moved early to the transit lounge to complete paperwork, receive medications, and undergo allied health reviews.
2. Enhanced Discharge Planning & Communication
  - Regular morning meetings between junior doctors and nursing teams to coordinate inpatient care and discharge plans.
  - Late afternoon planning meetings between the Senior Medical Officer (SMO) and Advanced Trainee Registrar to identify and discuss next-day discharges.
3. Earlier & More Efficient Ward Rounds & Meetings
  - Morning meetings (SMO, multidisciplinary team, journal club) to start at 0830.
  - SMO-led ward rounds to commence promptly at 0830 for timely decision-making.
4. Prioritizing Ward Duties for Junior Doctors
  - Attendance at multidisciplinary team meetings and journal clubs is no longer mandatory for junior doctors, allowing them to focus on ward tasks and discharge preparation, though attendance is encouraged when possible.

### Phase C

During the Baseline assessment period a total of 133 patients were discharged from the cardiology unit, with a median discharge time of 14:56. Of these, one (0.8%) patient was discharged before 10:00, 17 (12.8%) patients before 12:00, and the remaining after 12:00 (Table 2). In the period one month post intervention, compared to baseline, the median discharge time was 1h 27m earlier (95% confidence interval: 0h 11m – 2h 14m; p<0.01). Eight patients were discharged before 10:00, and 47 patients before 12:00, representing an 8- and 3-fold increase compared to 6 months pre-intervention, respectively. The results were similar in the period 6 months post intervention with median discharge time improving from baseline by 1h 20m (95% confidence interval: 0h 23m – 2h 10m; p<0.01). There was no significant difference in median discharge time between the one- and six-month post-intervention periods (p=0.42).

**Table 2.** Summary of numbers and proportions of patients discharged by certain times in the day, 6 months pre-intervention (Baseline), and 1-month and 6-months post intervention.

|  | **Total Patients discharged** | **Discharge before 10:00** | **Discharge before 12:00** | **Discharge after 12:00** |
| --- | --- | --- | --- | --- |
| Baseline | 139 | 1(0.8%) | 17(12.8%) | 121(87%) |
| One month post intervention | 127 | 8(6.3%) | 47(37.0%) | 72(56.7%) |
| Six months post intervention | 140 | 10(7.1%) | 41(29.3%) | 89(63.6%) |
| Total | 406 | 19(4.7%) | 105(25.9%) | 282(69.5%) |

**Figure 2.**
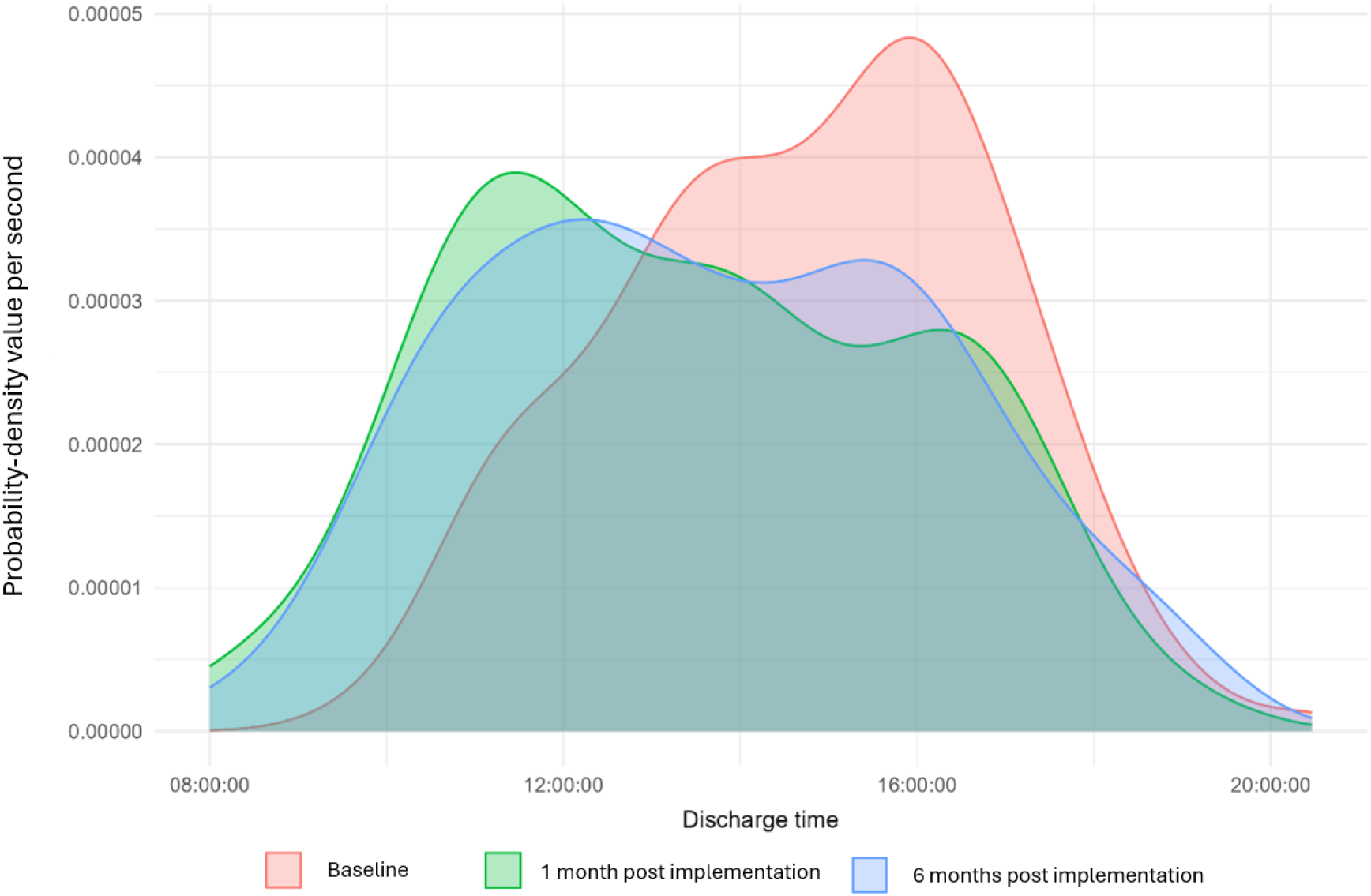
Density plot depicting mean time of discharge of patients at baseline, 1 month and 6 months post intervention.

We found that patients in both post-intervention groups were significantly more likely to discharge before 12:00pm (1 month: RR=3.03, 95% CI 1.84 – 4.99. p<0.01) (6 month: RR=2.39, 95% CI 1.43 – 4.01. p<0.01). Similarly, they were also more likely to discharge before 10:00 compared to patients in the Baseline group (1 month RR= 8.76, 95% CI 1.11 – 69.04. p=0.02) (6 month RR=9.93, 95% CI 1.29 – 76.52. p=0.01). Finally, there was no significant difference between 1 month vs 6 month groups regarding discharge before 10:00 (RR=1.13, 95% CI = 0.46 - 2.76, p=0.81) or discharge before 12:00pm (RR=0.78, 95% CI= 0.56 – 1.11, p=0.17).

Following successful results at the 6 month post-intervention period, we formalised the intervention as standard policy for the department.

## Discussion

Our study demonstrated that by using a Plan-Do-Check-Act model we were able to create a set of interventions which once implemented provided sustained improvements in discharge times. There was a significant increase in the number of patients discharged in the morning at both periods one and six months post intervention. Existing evidence suggests that a singular act of asking clinicians to prioritise seeing those patients suitable for discharge first on their ward round does not result in an earlier discharge time or a shorter length of stay [16]. Our results contrast with the study by Burden et al. which did not provide specific instructions for physicians on how to facilitate early discharges, nor did it consider specific departmental needs [16]. In comparison, in we created specific policies that were tailored to the circumstances of our department after analysing the root causes of delay – which we believe to be a major contributing factor to the success of our intervention.

We identified that a delayed ward round was one of the major factors contributing to late discharge. This was due to other patients taking priority or delayed commencement of ward rounds. As a result of teaching sessions and meetings, consultants arrived on the wards at inconsistent times triggering delays in the initialisation of the round. Late finishes to these meant resident medical officers were unable to begin the ward round until later in the morning. This created a knock-on effect with necessary reviews by other teams such as pharmacy being subsequently delayed. It also resulted in resident medical officers having less time to complete discharge summaries. Scheduling a regular daily meeting between medical and nursing team staff with the sole purpose of facilitating discharge planning was also crucial to earlier discharges. Delayed discharges are both a clinical risk and an economic burden to hospitals as they create bed shortages; patients who are unable to receive treatment have increased morbidity and mortality rates, and the opening of unfunded beds is costly [20]. We took a pragmatic approach regarding delayed reviews from other teams such as pharmacy and transferred patients to the hospital’s transit lounge, where they could be reviewed by allied health teams. Overall, implementation of our guidelines in Phase B saw a significant increase in the number of timely discharges, including an over two-fold increase in the number of discharges before noon. This is clinically significant as in our department by noon we would typically have several emergency department patients already waiting for a bed. These patients were then able to be transferred to the ward sooner, relieving bed blocking and ambulance ramping pressures. This is important especially due to the typical midday peak in presentations in Australian emergency departments and thus having beds available before then is crucial in alleviating patient load [4]. Additionally, there is an increasing recognition of the importance of discharges earlier in the morning due to the risks of late afternoon discharges which include increased mortality, infection rate and mood disturbances [5, 20-22].

In regard to past literature, Kirubarajan et al. suggests that there is no benefit to early discharge in relation to overall length of stay and mortality [23]. Their study has several limitations which must be considered. Firstly, it was a non-interventional retrospective study – we suggest it would be tenuous to thereby infer that earlier discharge times are conclusively unrelated to patient outcomes. A lack of inter-departmental communication meant a discharged patient’s bed was not made available immediately which could have influenced length of stay. Furthermore, it could be argued their results are not applicable to the Australian healthcare environment; between 2023-2024 across all states and territories, ED length of stay was 6h 05m in comparison to their median of approximately 14 hours [24].

Using the Plan-Do-Check-Act model and first investigating the root causes of discharge delay and then subsequently applying the knowledge into intervention planning, we were able to generate an effective and acceptable protocol, [17, 18]. As such, we hope our study will serve as an example which other hospital units can apply and utilise to reduce discharge times. Additionally, this study is the first of its kind in the context of a cardiology team and thus may be beneficial to other similar units due to similarities in patient discharge needs. We are also the first study to demonstrate the sustainability of such an intervention in an inpatient adult setting with encouraging results after 6 months highlighting the importance of adapting such guidelines to a local context such that it is more acceptable and feasible to staff.

### Limitations

One limitation of our study is that we did not collect any data on patient re-admission rates, which is often considered an important metric in gauging the effectiveness of early discharge; however, there is considerable evidence in the literature suggesting that early discharge does not increase re-admission rates if done safely and effectively [25-27]. The sample size for initial data collection regarding reasons for delayed discharge was quite small (n=20). However, the team conducting the study had considerable clinical experience collectively and felt that the vast majority of reasons for delayed discharge were covered and that a larger sample size was unlikely to have provided any additional barriers to discharge. Finally, the reasons we identified for delayed discharge were specific to our hospital and may not be broadly applicable. We would recommend other departments wishing to replicate our results utilise a similar approach using the Plan-Do-Check-Act methodology.

## Conclusion

We demonstrated that by applying a standardised quality improvement model (Plan-Do-Check-Act) in the context of a cardiology department, we were able to improve discharge times and the number of morning discharges. By identifying and addressing root causes of delayed discharges, we saw a notable reduction in average discharge time. Key interventions included prioritizing punctual morning rounds, optimising interdisciplinary communication, and effective utilisation of the transit lounge. Our findings highlight the effectiveness of a systematic approach to improving discharge processes, serving as a model for other similar hospital units seeking to optimize patient flow and hospital efficiency.

## Data Availability

All data produced in the present study are available upon reasonable request to the authors

## Acknowledgements & funding

The authors received no external funding for this research and therefore acknowledge no specific funding sources.

## Availability of data and materials

Datasets used and analysed during this study are available from the corresponding author upon reasonable request.

